# CD64 and CD169 could help differentiate bacterial from viral infections in Emergency Department

**DOI:** 10.1101/2020.10.28.20221259

**Authors:** Pénélope Bourgoin, Thomas Soliveres, Alexandra Barbaresi, Anderson Loundou, Isabelle Arnoux, Denis Bernot, Pierre-Emmanuel Morange, Pierre Michelet, Fabrice Malergue, Thibaut Markarian

## Abstract

**Background:** The identification of a bacterial, viral or even non-infectious cause is essential in the management of febrile syndrome in the emergency department (ED) setting, especially in epidemic contexts such as flu or CoVID-19.

**Objectives:** The aim of this study was to assess discriminative performances of two biomarkers, CD64 on neutrophils (nCD64) and CD169 on monocytes (mCD169), using a new flow cytometry procedure, in patients presenting with fever to the ED. Human leucocyte antigen-DR on monocytes (mHLA-DR), HLA-ABC ratio (rHLA-ABC), and CD64 on monocytes (mCD64) were also assessed.

**Methods:** 85 adult patients presenting with potential infection were included during the 2019 flu season in the ED of La Timone Hospital. They were divided into four diagnostic outcomes according to their clinical records: no-infection, bacterial infection, viral infection and co-infection.

**Results:** mCD169 was elevated in patients suffering from *Flu A* virus or Respiratory Syncytial Virus, while nCD64 was mainly found elevated in subjects with *Streptococcus pneumoniae*. In total, 38 (45%) patients were diagnosed with bacterial infections, 11 (13%) with viral infections and 29 (34%) with co-infections. nCD64 and mCD169 showed 90% and 80% sensitivity, and 78% and 91% specificity, respectively, for identifying patients with bacterial or viral infections. Other biomarkers had lower discriminative performances.

**Conclusions:** nCD64 and mCD169 have potential for accurately distinguishing bacterial and acute viral infections. Combined in an easy and rapid flow cytometry procedure, they constitute a potential improvement for infection management in the ED setting, and could even help for the triage of patients during emerging epidemics.

## 1. INTRODUCTION

In the management of febrile syndrome, diagnostic guidance toward an infectious etiology is essential (1). The characterization of a viral, bacterial or other infectious or non-infectious cause allows early appropriate patient management. Nevertheless, the data in the literature report the complexity of triage and diagnostic guidance (2). The integration of the clinical examination is essential, but unfortunately is often not enough (3); diagnostic elements are therefore needed as quickly as possible in the emergency department (ED), especially in epidemic contexts such as flu or CoVID-19 (4–6).

Biological markers such as procalcitonin (PCT) and C-reactive protein (CRP) are commonly used in clinical practice. These markers have bacterial specificity but share a wide range of values with viral infections and do not make it possible to exclude or to confirm definitively the diagnosis (7–9). Their triage capacity in ED is thus regularly challenged (10). Measuring multiple specific biomarkers simultaneously, with a simple technique and rapid time-to-results, would be better compatible with the needs of triage in emergency medicine (11).

We developed a rapid flow cytometry assay, able to measure leucocytes biomarkers expressions within 10 minutes (12), and demonstrated promising results for the triage of patients with fever at the Emergency Department (13,14), with CD64 on neutrophils (nCD64), increased in case of bacterial infections (15), and CD169 on monocytes (mCD169), increased in case of viral infections (16). In these previous studies, one limitation was the low number of infected patients, especially those with viral diseases.

In this new study, we thus included more patients, and focused on those with infectious symptoms during the flu season. The main goal was to confirm the relevance of CD64 and CD169 for discriminating between bacterial and viral infections in such an epidemic context.

Secondary goal was to assess the relevance of other interesting biomarkers, including (13): Human Leucocyte Antigen (HLA)-DR on monocytes (mHLA-DR), HLA-ABC ratio (rHLA-ABC) of monocytes/neutrophils and CD64 on monocytes (mCD64).

## 2. METHODS

### 2.1. Studied population

The study population included patients older than 18 years, if they presented to the adult ED of La Timone University Hospital in Marseille, France, with infection signs, during 2019 flu season. The inclusion criteria were the presence of fever greater than 38°C or hypothermia less than 36.5°C, and any potential respiratory (cough, sputum, and dyspnea), urinary (potential urinary infection), abdominal (pain syndrome, diarrhea), cutaneous (erysipelas), or neurological (meningitis) infectious clinical signs. The exclusion criteria were: incomplete clinical files, traumatized patients or patients presenting with a known inflammatory or autoimmune disease, neoplasia, chronic infectious disease (viral, fungal, or bacterial), or antibiotic, antiviral, or immunosuppressive treatment prior to admission, and patients with extensive burns or recent surgery (less than 1 month).

Their routine care was not modified, and confidentiality was preserved at all levels. All enrolled patients provided informed consent and no objection authorization, so that their data could be retrieved from their clinical records by a team of emergency department specialists, and could be used in the study.

This observational and noninterventional prospective study was approved by the La Timone Hospital Ethical Committee and the Committee for Protection of Persons (CPP approval no. 181160; ID-RCB approval no. 2018 A02706-49). Procedures followed were in accordance with the Helsinki Declaration.

### 2.2. Clinical data collection

Electronic medical records were retrieved for each patient by a team of ED specialists:

- epidemiological data: sex, age, clinical history (evolutionary cancer, liver disease, congestive heart failure, cerebrovascular disease), medical institutionalization, altered mental status;
- physiological data: cardiac parameters (systolic, diastolic and average blood pressures, pulse and respiratory rates), body temperature, fever duration, vital signs and symptoms (respiratory, abdominal, neurological, urinary, cutaneous or others), and eventual oxygen-, antibiotic- or antiviral-therapy;
- clinical data: time from onset, symptoms, X-ray examination results (performed and atypical chest X-ray or ultrasound or CT scan), final diagnosis established by the ED practitioner, outcome of the ED visit (released home, conventional or critical care hospitalization), and eventually duration of the hospitalization;
- and biological data: white blood cell (WBC) and polymorphonuclear neutrophil (PMN) counts, CRP and PCT levels, biochemical measurements (urea, sodium, glucose, hematocrit, hemoglobin), and name of the identified pathogens if isolated.

WBC and PMN counts were assessed using a Sysmex XN system (Sysmex Inc., Kobe, Japan). PCT was measured using a Dosage ADVIA Centaur BRAHMS Procalcitonin system (Siemens, Munich, Germany) and CRP using Gen.3 system (COBAS, Roche, Basel, Switzerland).

Isolation of potential viruses relied on examining blood or cerebrospinal fluid samples with polymerase chain reaction (PCR), and serum IgG and IgM with LIAISON analyzer (DiaSorin, Saluggia, Italy). Potential bacteria were detected in blood and respiratory tract secretion (nasopharyngeal swabs, tracheobronchial aspirate, or bronchoalveolar lavage) cultures by a Bruker Mass Spectrometry system (Brucker Inc., Billerica, MA, USA), and in urine cultures by urinary antigen tests (for *Pneumococcus* and *Legionella*) or PCR (for *Chlamydia* and *Mycoplasma*).

### 2.3. Adjudication committee

Based on all clinical and biological data, an adjudication committee classified patients in 4 groups.

Group I: Subjects for which infection was ruled out. No evidence of infection was found according to clinical symptoms and laboratory test results.

Group II: Subjects diagnosed with bacterial infections. Subjects were categorized into this group either on the basis of a positive bacterial culture result, and/or if clinical and laboratory findings such as concomitant high levels of CRP and PCT and negative viral test results strongly suggested the presence of a bacterial infection.

Group III: Subjects diagnosed as having viral infections. This group contained subjects that presented with typical clinical symptoms of infections, but negative bacteriological results and/or low PCT levels. In some cases, viral agents were found by antigen-based tests or serological assays.

Group IV: Subjects diagnosed as having both viral and bacterial infections. The committee was not aware of the flow cytometry results.

### 2.3. Flow cytometry testing

Leftover ethylenediaminetetraacetic acid (EDTA)-treated blood samples were pseudonymized, and processed by flow cytometry according to a newly described one-step procedure (12). Briefly, a multicolor panel was constituted and dried as a “glassified” layer at the bottom of a 5-mL testing tube using the DURA Innovations drying process (Beckman Coulter Inc.), with antibodies at their optimized amounts for a single test: anti-CD169-phycoerythrin (PE) (clone 7-239), anti-CD64-PacificBlue (PBE) (clone 22), anti-HLA-DR-allophycocyanin (APC) (clone Immu357) and anti-HLA-ABC-Alexa Fluor 700 (AF700) (clone B9.12.1), all custom products from Beckman Coulter Inc. (Brea, CA, USA).

For each blood sample tested, 500 μL of Versalyse lysing solution (Beckman Coulter Inc.) and 5 μL of EDTA-treated blood were transferred to one dried tube. After incubation for 15 minutes, samples were analyzed on a three-laser, 10-color Navios flow cytometer (Beckman Coulter Inc.). Flow-Set beads (Beckman Coulter) were used before each analytical run in order to control the variability in device performance, however no harmonization between the measured values over the study period was necessary. Analysis was performed using Kaluza Analysis Software (version 2.1; Beckman Coulter Inc.).

Leucocytes were gated using Side Scatter (SSC) and CD64 expressions, as lymphocytes (low SSC, CD64-), monocytes (intermediate SSC, CD64+) and neutrophils (high SSC), prior to the analysis of nCD64, mCD169, mHLA-DR, rHLA-ABC and mCD64. Results were expressed as mean of fluorescence intensities (MFI).

### 2.4. Statistical analysis

Statistical analysis was performed using IBM SPSS Statistics version 20 (IBM SPSS Inc., Chicago, IL, USA). Quantitative data were expressed as mean ± standard deviation. Qualitative variables were expressed as frequency with percent.

Comparisons of quantitative variables among the different groups were performed using Student’s t-test or Mann-Whitney U test. Comparisons of percentage were performed using Khi-2 or Fisher’s exact tests if conditions were missing. Comparisons of more than two groups were performed by Freeman-Halton extension of Fisher’s exact test for qualitative variables and by analysis of variance or Kruskall-Wallis tests for quantitative variables.

The ability of biomarker levels to discriminate between bacterial and viral infections was investigated by means of receiver operating characteristic (ROC) curve analysis. ROC curves were used to define the best threshold of biomarker indexes. Analyses were based on area under the curve (AUC), sensitivity (true positives / positives [TP / P]), specificity (true negatives / negatives [TN/N]), positive likelihood ratio (sensitivity / [100 – specificity]) and negative likelihood ratio ([100 – sensitivity] / specificity]). All values were expressed as ranges (between 0 and 100), with 95% confidence intervals. For all tests, two-sided *p* values less than 0.05 were considered statistically significant.

## 3. RESULTS

### 3.1. Clinical features of the patients

During the study period, 104 patients admitted to the ED of La Timone Hospital were included. All blood samples were processed by flow cytometry, but only 85 out of the 104 subjects satisfied the inclusion criteria. Nineteen patients for whom clinical files were incomplete, and in particular for whom there was doubt about the presence of underlying or asymptomatic infections that were not sufficiently documented, were removed from the study, because it could have biased their final diagnosis and classification.

The adjudication committee classified the 85 remaining patients: 7 (8%) were defined as not-infected, 38 (45%) as bacterially infected, 11 (13%) as virally infected and 29 (34%) as presenting with both a bacterial and a viral infection (co-infection). An overview of the study workflow is shown in Figure 1.

**Figure 1.**
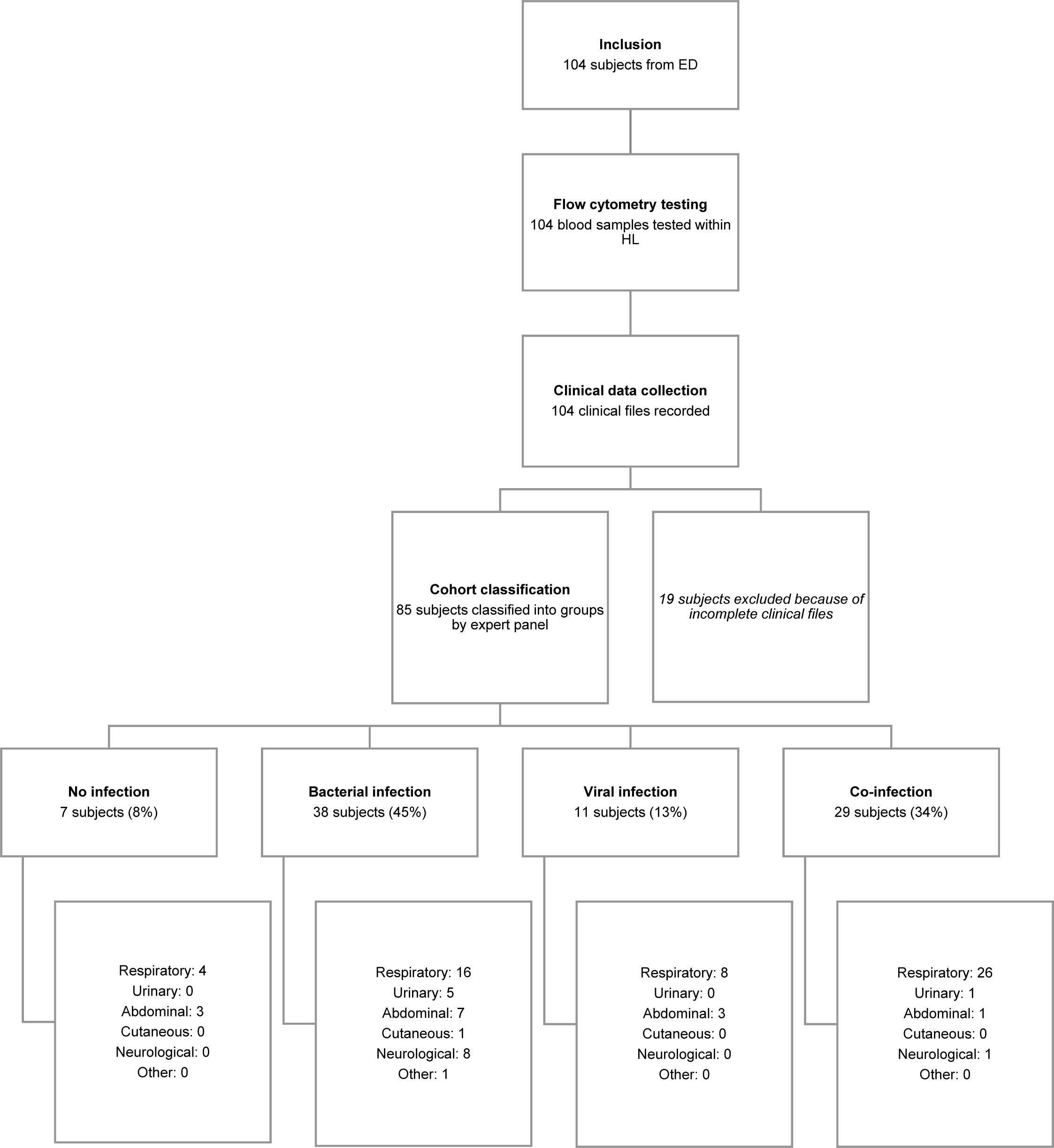
Overview of the study workflow. Representation of each step of the study, conducted between the Emergency Department (ED) and the Hematology Laboratory (HL) of La Timone Hospital, with final numbers of included and excluded subjects, and details about clinical kinds of symptoms.

### 3.2. Clinical epidemiology and biomarker levels

The final cohort consisted of 85 patients, including 30 (35%) women and 55 (65%) men, with a mean age of 56 (± 25) years. Their epidemiological, clinical, and biological data are presented in Table 1. Bacterial infections were considered as patients presenting with bacterial infections only plus bacterial co-infections, whereas viral infections were considered as patients presenting with viral infections only plus viral co-infections.

**Table 1.**
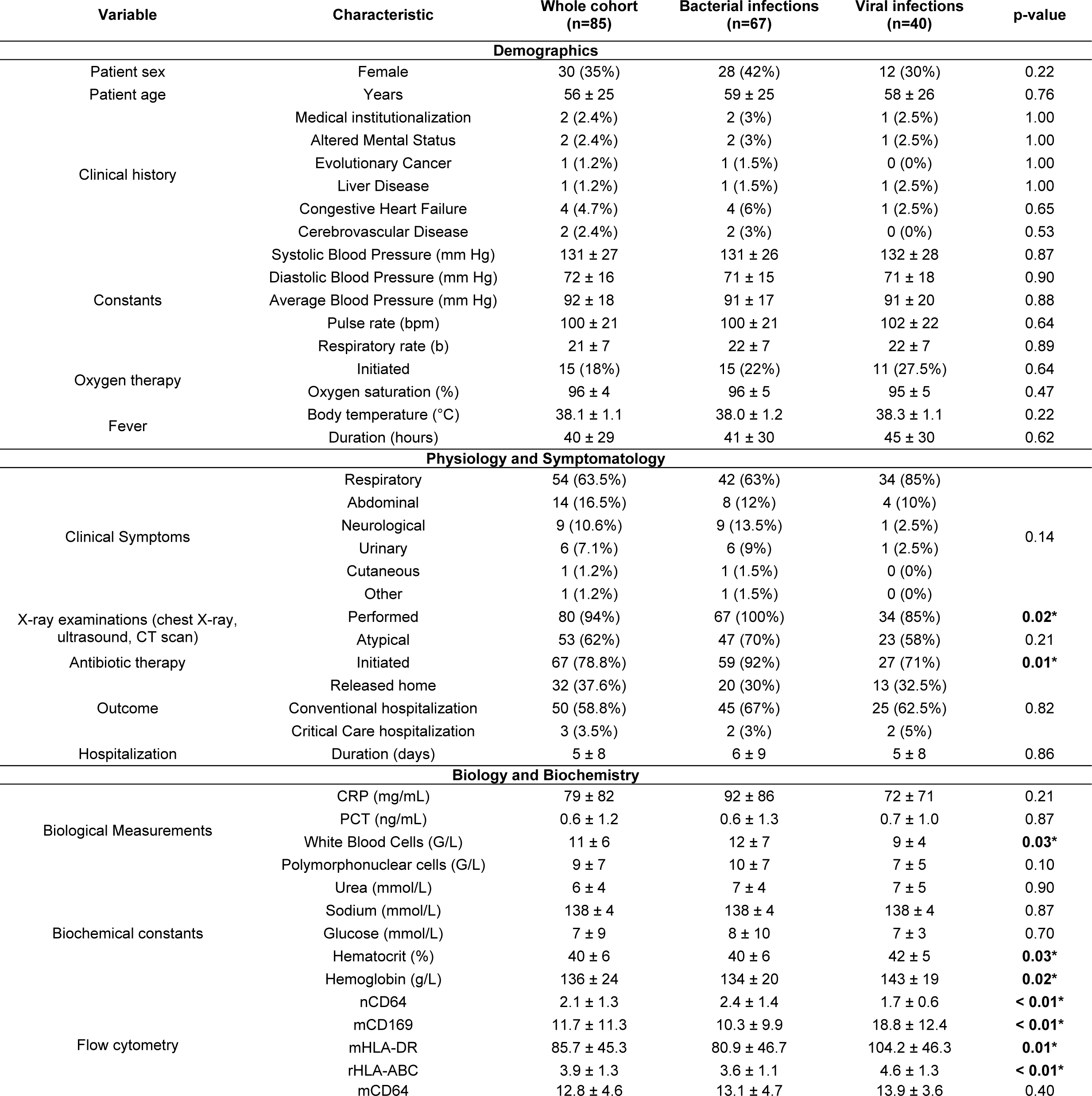
Cohort study characteristics. Clinical and biological data for the whole cohort and bacterially and virally infected subjects. Values are presented either as mean ± standard deviation or as number with percentage. Bacterial infections are defined as bacterially-infected plus bacterially-co-infected subjects, whereas viral infections are defined as virally-infected plus virally-co-infected subjects. Variables for which p values were less than α = 5% are indicated in bold with * to indicate statistically significant differences between both groups.

No significant differences between bacterial versus viral infection groups were observed for gender ratio (*p: 0.22*), mean age (*p: 0.76*), clinical histories and features of the patients. Overall, patients had a mean elevated body temperature (38.1 ± 1.1°C), and temperature was not significantly different between bacterial versus viral infection groups (*p: 0.22*).

A wide range of infectious symptoms was observed among groups (respiratory, abdominal, neurological, urinary, cutaneous, and other). The most common clinical presentation associated with bacterial (n=42; 63%) and viral (n=34; 85%) infections was respiratory. As expected, based on symptoms, patients diagnosed as having bacterial infections were treated more frequently with antibiotics (n=59; 92%; *p: 0.01*), and X-ray examinations were performed more frequently (n=67; 100%; *p:* 0.02). After their ED medical consultation, 32 patients (38%) could have been released home, but 53 patients (62%) were admitted to hospital specialized departments or the critical care department for longer observation. However, all patients that were kept at the hospital for observation remained for a non-significantly different duration (*p*: *0.86*).

Finally, comparison of biochemical and biological measurements showed significant differences between bacterial versus viral infection groups for WBC count (*p: 0.03*), hematocrit (*p: 0.03*) and hemoglobin (*p: 0.02*) levels. PCT (*p: 0.87*) and CRP (*p: 0.21*) were not significantly different among bacterial versus viral infection groups.

Overall, 94 common pathogen species were detected (Table 2). The most frequent pathogens were *Streptococcus pneumoniae* (38%) and *Flu A virus* (26%).

**Table 2.**
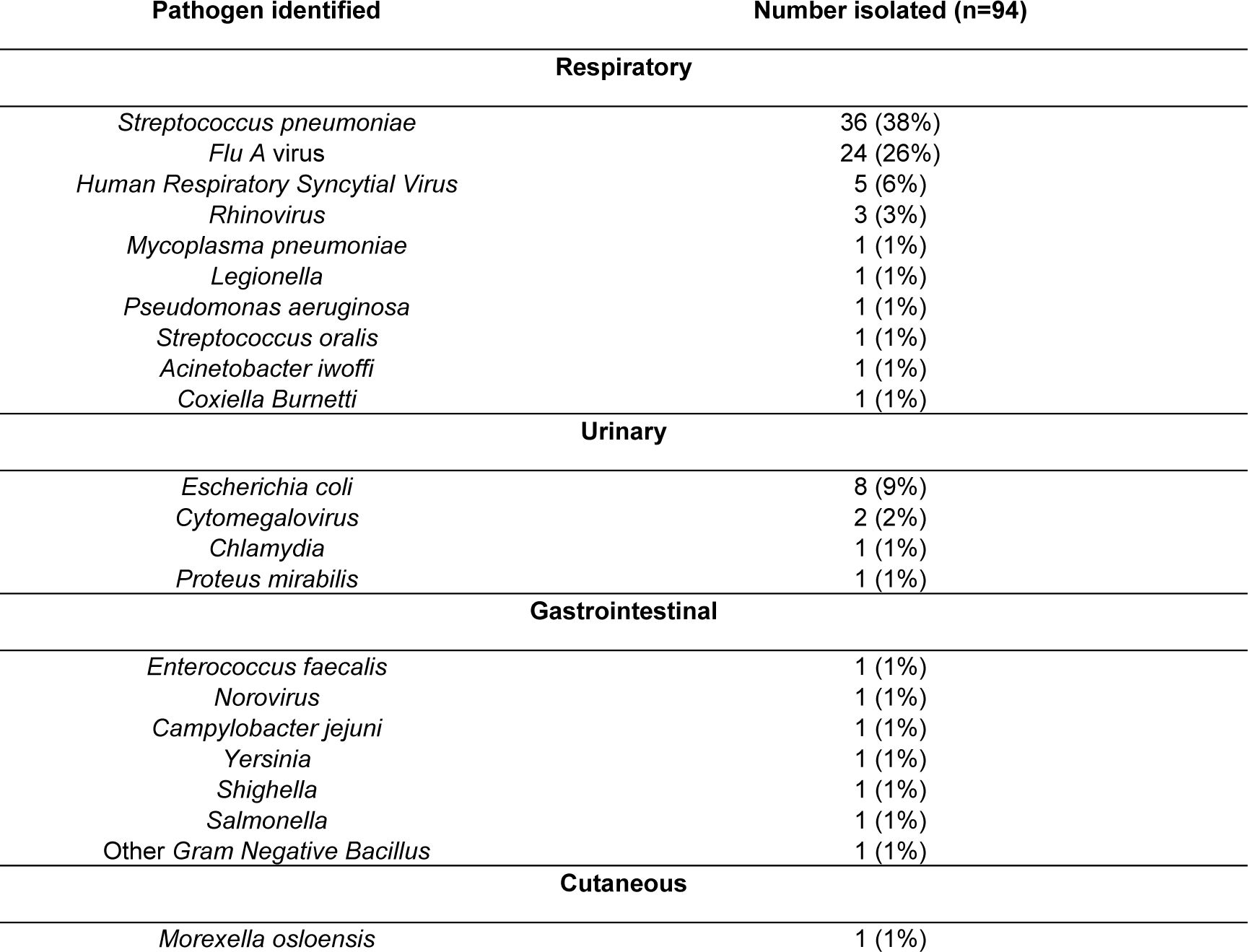
Identification of pathogens in the cohort. Name and number (percentage in brackets) of isolated bacteria and viruses in the whole cohort.

Patients with bacterial infections had higher nCD64 (MFI of 2.4 ± 1.4; *p < 0.01*) and lower mHLA-DR (MFI of 80.9 ± 46.7; *p: 0.01)* levels. Conversely, patients with viral infections had higher mCD169 (MFI of 18.8 ± 12.4; *p < 0.01*), mHLA-DR (MFI of 104.2 ± 46.3; *p*: *0.01*) and rHLA-ABC (MFI of 4.6 ± 1.3; *p < 0.01*) levels, when compared among bacterial versus viral infection groups. Finally, mCD64 levels were not significantly different among these groups (*p: 0.399*). Example of flow cytometry results is shown in figure 2.

**Figure 2.**
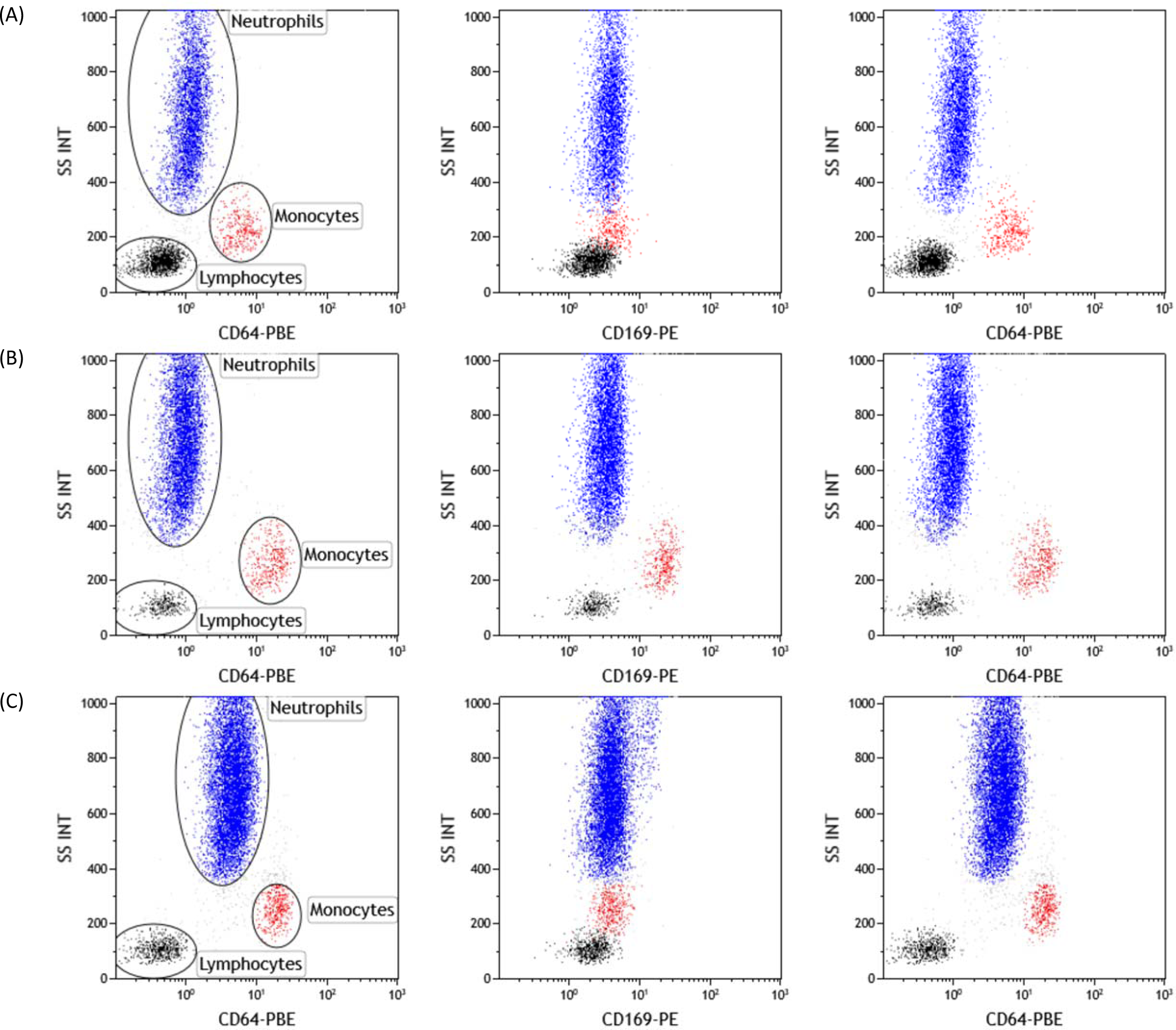
Biomarkers expressions in whole blood. Leucocytes were gated using Side Scatter (SSC) and CD64 expressions, as lymphocytes (low SSC, CD64-; in black), monocytes (intermediate SSC, CD64+; in red) and neutrophils (high SSC; in blue). Examples of CD169 and CD64 expressions on leucocytes were given for 3 subjects: (A) one first healthy volunteer whole blood, (B) one second viral-infected whole blood, and (C) one third bacterial infected whole blood.

### 3.3. Biomarker ROC analysis

ROC analysis of the five biomarkers was made for evaluating their performance to identify the bacterial or viral etiology of an infection (Figure 3).

**Figure 3.**
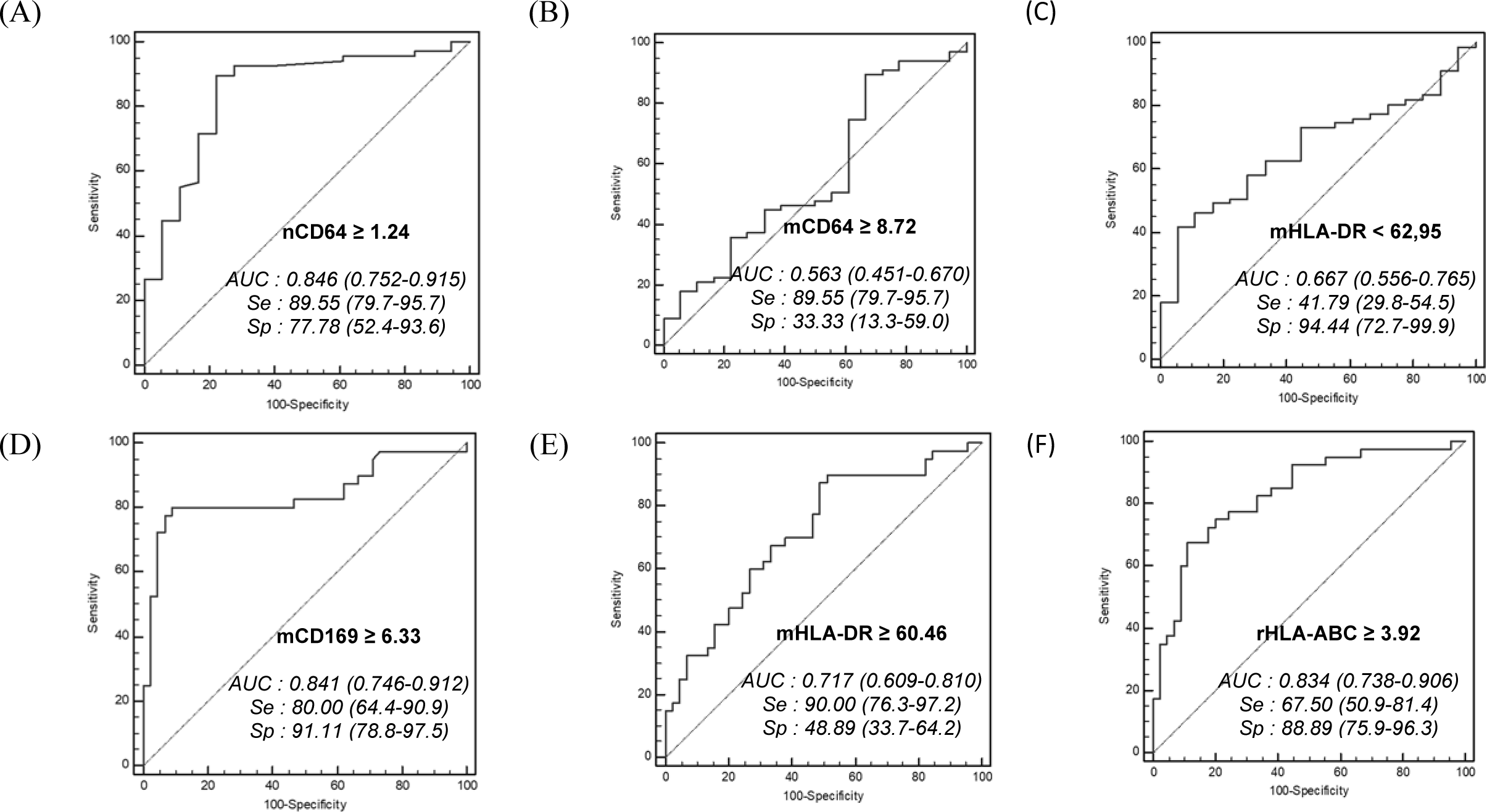
ROC analysis. ROC curves for the discrimination of bacterial infections with (A) nCD64, (B) mCD64 and (C) mHLA-DR. ROC curves for the discrimination of viral infections with (D) mCD169, (E) mHLA-DR and (F) rHLA-ABC. Optimal thresholds for each biomarker are indicated with calculated values of specificity (Sp) and sensitivity (Se). Area under the curve (AUC) and 95% confidence interval are also given for each ROC analysis.

Using a cutoff point of greater than or equal to 1.24 for patients with bacterial infections, the results indicated that the nCD64 MFI showed the best bacterial discriminative performance, with a sensitivity of 90% (80–96%), specificity of 78% (52–94%), positive likelihood ratio of 4.03 (1.7–9.6), and negative likelihood ratio of 0.13 (0.06–0.3). The area under the curve was of 0.85 (0.75–0.92).

Using a cutoff point of greater than or equal to 6.33 for patients with viral infections, the results showed that the mCD169 MFI exhibited the best viral discrimination, with a sensitivity of 80% (64–91%), specificity of 91% (79–98%), positive likelihood ratio of 9.00 (3.5–23.2), and negative likelihood ratio of 0.22 (0.1–0.4). The area under the curve was of 0.84 (0.75–0.91).

ROC analysis of the three other biomarkers was more equivocal. To detect bacterial infection, using a cut-off-point greater than or equal to 8.72, mCD64 MFI exhibited good sensitivity of 90% (80-96%) but lower specificity of 33% (13-59%). Conversely, using a cut-off point of less than 63.0, mHLA-DR MFI exhibited good specificity of 94% (73-100%) but low sensitivity of 42% (30-55%).

To detect viral infections, using a cut-off-point greater than or equal to 60.46, mHLA-DR MFI exhibited good sensitivity of 90% (76-97%) and low specificity of 49% (34-64%), whereas, using a cut-off-point greater than or equal to 3.92, rHLA-ABC exhibited low sensitivity of 68% (51-81%) but good specificity of 89% (76-96%).

## 4. DISCUSSION

This study aimed to investigate the value of nCD64 and mCD169 biomarkers in a population of patients with febrile symptoms in a clinical context of the ED, using a new procedure of flow cytometry. Assessment of their levels, in non-infected, bacterially-infected, virally-infected or co-infected subjects, demonstrated, within this study, their relevance for identifying etiology of infections.

In this new study, a total of 85 subjects were included, and divided into four diagnostic outcomes: not-, bacterially-, virally-or co-infected subjects. Among these groups, each outcome was always represented by more than 10 subjects. Moreover, almost all cases were confirmed by a bacterial or viral isolate in biological samples (22 different pathogens found among 94 isolates). As patients were included during flu season, the most frequent pathogens were respiratory: *Streptococcus pneumoniae, Flu A* virus *and Human Respiratory Syncytial Virus*. These pathogens are commonly expected during this period, as has been reported (5).

CD64 on neutrophils and CD169 on monocytes were the two main biomarkers assessed in this study by flow cytometry for discriminating between bacterial versus viral infections. Expressions of these two biomarkers have been shown to be directly induced, within hours, by interferons produced by the body in response to pathogen detection (14). Ability of nCD64 to discriminate between bacterial and non-bacterial infections has been largely demonstrated for years (17), whereas mCD169 increase after infection by viruses has only been described recently. mCD169 seems to be a general biomarker of acute viral infections since it has been found in patients with HIV (16,18,19), EBV (20), RSV (21), CMV (22), Dengue (23,24), Zika (25), noroviruses (26), Lassa and Marburg (27). Here, high levels of mCD169 have been observed for the first time for *Flu A* virus.

High levels of sensitivity and specificity were found in the study for both biomarkers. Interestingly, nCD64 showed a better sensitivity of 90% than specificity of 78%, whereas mCD169 showed a better specificity of 91% than sensitivity of 80%. These results further demonstrate their valuable use for infection etiology guidance in ED settings for patient triage. Indeed, a biomarker used for bacterial infection identification in ED needs to be as sensitive as possible to detect the majority of cases, demonstrating at least 90% sensitivity, as any missing case could delay patient from receiving appropriate antibiotic therapy, and thus increase their risk of developing sepsis and progression to death. Conversely, a viral marker in ED has to be very specific to ensure the etiology, with at least 90% specificity, as it allows the practitioner to discharge the patient, as well as avoiding the empirical use of antimicrobial drugs in cases where they are not required (28).

The global health issue of overuse of antibiotics has been illustrated in this study: the epidemiological data showed that an antibiotic therapy was initiated in 67 out of 85 patients. Examining the whole cohort, 36 out of 38 subjects with bacterial infections, and 23 out of 29 co-infected with bacteria and viruses, appropriately received antibiotics. Conversely, 4 out of the 11 subjects virally infected only, and 4 out 7 not-infected subjects, received antibiotics, cases where antibiotics are ineffective, or even worse, might be dangerous (29). As a consequence of their usefulness for determining etiology, both biomarkers could be incorporated as part of the overall clinical management of patients with fever, and used for evaluation of antibiotic therapy initiation, duration and end (30).

Three other secondary biomarkers were evaluated in the study: HLA-DR on monocytes, HLA-ABC ratio of monocytes/neutrophils and CD64 on monocytes. The first two biomarkers have been identified among 13 other leucocyte biomarkers as being the most promising for determining infectious etiology (13), and the third has been considered in some studies for comparison to nCD64. Firstly, HLA-DR is widely known to be decreased in sepsis subjects (31), but also, as it is the major histocompatibility class II, it has also been seen to increase in cases of viral infection (13). Secondly, rHLA-ABC, although less frequently evaluated, normally increases in cases of viral infection (32). Finally, mCD64 is known to evolve similarly as nCD64, by increasing in cases of bacterial infection, although demonstrating lower performance (33). These three biomarkers were evaluated in this study to clarify and confirm their promising use for infectious etiology determination. Unfortunately, they did not show expected performances: either their sensitivity or specificity was not sufficient enough to ensure effective identification of bacterial or viral infections.

Finally, one major issue remaining is how to measure the biomarkers in ED clinical practice. The ED environment is complex and dynamic, and thus requires technologies tailored specifically for prevention, diagnosis, and outcome of infection, to enhance patient safety in emergency care. A new solution was demonstrated by assessing the levels of expression of the markers using an innovative, 15-minute, one-step method of flow cytometry (12). This procedure may meet the minimum characteristics required for a bacterial versus viral bedside test, as targeted by Dittrich team (11): 1) the use of a capillary blood drop could be potentially used as only 5 µL are necessary, although EDTA blood samples were used here; 2) reagent storage and laboratory procedures are carried out at room temperature; and 3) all steps for testing the samples are combined into one step, with no specific material or training needed. In summary, this assay yields results in less than 15 minutes from initial blood collection, with discriminative power greater than other currently existing tests. The procedure used here however, for the moment has limitations regarding the instrument: on one hand, MFI measurements would necessitate standardization efforts to avoid variations due to instrument setup, and on the other hand, flow cytometer sample preparation and analysis would need automation.

The study has other limitations. First, patients were enrolled from the ED during approximately 5 months, and only within one ED from one hospital. Even if a good representation of the most common infections was achieved, it is not sufficient to provide a complete validation of biomarkers. In future studies, it might be preferred to extend the study to other ED from other hospitals. Secondly, biomarkers seemed to be useful for infected patients, but their kinetics are not well known. It is important to understand their delay of onset after infection and their sequential evolution in blood circulation. Future research should thus focus on measuring these biomarkers on sequential samples from same subjects, and evaluating their prognostic value, both for bacterial versus viral infection diagnosis and for therapy duration.

## 5. CONCLUSION

In summary, CD64 and CD169 were confirmed to be biomarkers of interest to predict bacterial versus viral infection causes of fever, whereas HLA-DR and HLA-ABC demonstrated lower performances in these settings. Flow cytometry is currently the universally applied method for identifying cell surface markers, but in this context, its availability remains limited in emergency settings. As part of a global effort to reduce inappropriate antibiotic use, this study makes available the measurement of infection-related biomarkers using a new flow cytometry procedure, promisingly applicable at the point-of-care. This association of infection related biomarkers and flow cytometry is promising to facilitate an easy, rapid and robust discrimination of bacterial versus viral infections and thus the successful care of patients with potential infection presenting to the Emergency Department.

## Data Availability

The authors certify that this manuscript reports original clinical research data. Individual data that underlie the results reported in this article are available from the corresponding author following publication, including the study report and study protocol. Additional data are available upon reasonable request.

## ETHICS AND PATIENT APPROVAL STATEMENT

The authors state that they have followed the principles outlined in the Declaration of Helsinki for all human experimental investigations. Routine care of the subjects was not modified; analyses were performed on anonymized blood left over, and all data collected in the study were part of routine clinical practice and retrieved from subject records. Results of the study had no influence on subjects’ management.

This observational and noninterventional prospective study was approved by the La Timone Hospital Ethical Committee and the Committee for Protection of Persons (CPP approval no. 181160; ID-RCB approval no. 2018 A02706-49).

## DECLARATION OF INTEREST

Fabrice Malergue and Pénélope Bourgoin are Beckman Coulter employees.

## FUNDING SOURCE

Pénélope Bourgoin is recipient of a CIFRE PhD grant (n°2016/1368) from the ANRT (National Agency for Research and Technology). This study was partially supported by Beckman Coulter through donations of the research reagents used in the study and participation of the two employees mentioned above. This private company had no role in the study design, or collection and interpretation of the clinical data. Beckman Coulter and the Beckman Coulter product and service marks mentioned herein are trademarks or registered trademarks of Beckman Coulter, Inc. in the United States of America and other countries. All other trademarks are the property of their respective owners.

## ACKNOWLEDGEMENTS

The authors would like to thank the technician team from the La Timone University Hospital Hematology Laboratory for their assistance in blood testing, the ANRT for funding the PhD grant, and Beckman Coulter for giving reagents used in the study.

## Notes

### Competing Interest Statement

Fabrice Malergue and Penelope Bourgoin are Beckman Coulter employees. The other authors declare no competing interest.

### Clinical Trial

NCT03912870

### Funding Statement

Penelope Bourgoin is recipient of a CIFRE PhD grant (2016/1368) from the ANRT (National Agency for Research and Technology). This study was partially supported by Beckman Coulter through donations of the research reagents used in the study and participation of the two employees mentioned above. This private company had no role in the study design, or collection and interpretation of the clinical data. Beckman Coulter and the Beckman Coulter product and service marks mentioned herein are trademarks or registered trademarks of Beckman Coulter, Inc. in the United States of America and other countries. All other trademarks are the property of their respective owners.

